## Supplementary document 1: Methods and materials for "The impact of climate suitability, urbanisation, and connectivity on the expansion of dengue in 21st century Brazil"

### S1 Document: Methods and material

#### Dengue surveillance and outbreak definitions in Brazil

Monthly dengue case data are freely available from Brazil's Notifiable Diseases Information System (SINAN), via the Health Information Department, DATASUS (http://www2.datasus.gov.br/DATASUS/index.php?area=0203). Although notification of a suspected dengue case is mandatory in Brazil, the surveillance system is predominantly passive, which means that many mild and asymptomatic cases may be missed. One investigation of the Brazilian dengue surveillance system estimated that there were 12 actual infections per reported case overall, which rose to over 17 in periods of high incidence (1). Rather than use dengue case data which differs in accuracy between regions, and between epidemic and non-epidemic periods, we aggregated the cases by year and converted them into a binary outbreak indicator where cases exceeded some outbreak threshold. Several methods have been used to define outbreak thresholds Brazil, including a monthly moving average, where historical data are used to estimate the expected number of cases within a region (2,3), and a fixed threshold based on the dengue incidence rate, defined by the Brazilian Ministry of Health as the number of cases per 100,000 residents (4). The fixed threshold approach was chosen as the monthly moving average set very low thresholds (sometimes less than one case) in parts of the country previously protected from dengue transmission. The annual dengue incidence rate was calculated using estimates of the annual population for each municipality obtained from the Brazilian Institute of Statistics and Geography (IBGE) via DATASUS (http://tabnet.datasus.gov.br/cgi/deftohtm.exe?ibge/cnv/poptbr.def). Our primary analysis used an outbreak threshold of more than 300 cases per 100,000 residents, defined as 'high risk' by the Brazilian Ministry of Health. We also tested a 'medium risk' indicator, defined as more than 100 cases per 100,000 residents (4).

#### Socioeconomic factors

We obtained information about the percentage of residents in each municipality living in urban areas, the percentage with access to the piped water system, and the percentage that had refuse collected (either privately or using the municipal service) from the 2000 and 2010 censuses via DATASUS. Despite Brazil having the largest economy in South America, it has been the most unequal since 2015 (5). Access to basic services differs greatly across the country and the traditionally wealthier regions in the South and Southeast have almost universal coverage at the municipality level in contrast to rural parts of the North and Northeast which had little or no access, even in 2010. We found that the level of urbanisation was highly correlated to access to piped water (Figure S5, r = 0.656, 95% confidence interval: [0.641, 0.671], p < 0.001) and refuse collection (Figure S5, r = 0.794, 95% confidence interval: [0.784, 0.804] p < 0.001) when aggregated to the municipality level. Therefore, access to piped water and refuse collection were not included in the models as they were not useful at explaining the differences within cities at this level of aggregation and would likely introduce multicollinearity into the model.

#### Hierarchical levels of influence of cities

We extracted the level of influence of cities from the Regions of Influence of Cities (“Regiões de Influência das Cidades”, REGIC) studies carried out by IBGE in 2007 and 2018 (6,7) to use as a proxy for human movement within our models. REGIC aims to recreate the complex urban network of Brazil using information from surveys about the frequency and reasons for the movement of people and goods around the country. The level of influence assigned to each city was based on the number of people travelling to the city but also the number of important institutions that attracts the movement of people from outside the city, such as hospitals, universities, business centres, government agencies, and cultural centres (such as theatres and shopping centres). Cities were classified into five levels:

1. Metropolis: the largest cities in Brazil, with strong connections throughout the entire country. This includes São Paulo, the capital Brasilia, and Rio de Janeiro.
2. Regional capital: large cities which are connected throughout the region in which they are located and to metropoles. This includes state capitals that were not classified as metropoles, such as Rio Branco, Campo Grande and Porto Velho.
3. Sub-regional capital: cities with a lower level of connectivity, mostly connected locally and to the three largest metropoles.
4. Zone centre: smaller cities with influences restricted to their immediate area, often neighbours.
5. Local centre: the smallest cities in the network which typically only serve residents of the municipality and are not connected elsewhere.

There were 12 metropoles, consisting of 203 municipalities, according to the 2007 REGIC study: São Paulo, Rio de Janeiro, Brasilia, Manaus, Belém, Fortaleza, Recife, Salvador, Belo Horizonte, Curitiba, Goiânia and Porto Alegre. In 2018, this increased to 15 metropoles, consisting of 214 municipalities, as Campinas, Florianópolis and Vitória were re-classified from regional capitals to metropoles. The number of regional capitals and sub-regional centres also increased between 2007 and 2018 from 70 to 97 and from 169 to 352 respectively. The number of lower-level cities, zone centre and local centres, both decreased from 556 to 398, and from 4473 to 4037 (Table S1). The distribution of highly connected urban centres is uneven across the country; the South and Southeast regions are particularly well connected, while the North and Northeast contain fewer high-level centres (Figure 3, Table S1). The proportion of higher-level centres has increased in each region of Brazil, although the Amazon rainforest remains less connected than other areas (Figure S6). Metropoles, regional capitals and sub-regional centres had higher levels of urbanisation, access piped water and refuse collection on average than less connected centres (Figure S7).

#### Modelling approach

We formulated a spatio-temporal generalised additive model (GAM) to quantify the relationship between temperature suitability, level of connectivity and socioeconomic conditions on the odds of a municipality experiencing an outbreak. The response variable was a binary outbreak indicator defined as an annual dengue incidence rate of more than 300 cases per 100,000 residents. To account for spatial and temporal patterns in the data, smooth functions of the year and the coordinates of the centroids of municipalities were included in the model. We used thin plate regression splines to represent the smooth (2D) function of the coordinates. Thin plate splines are data-driven and estimate the best fitting function for the data (8). To account for changes in spatial patterns over the period, we also included a space-time interaction term created by applying a tensor product smooth to the coordinates and the year. Tensor product smooths allow interactions between variables that are measured on different scales (in this case, space and time). The final model equation was as follows:

$$Y_{it}\sim Bernoulli(p_{it})$$

$$logit(p_{it})=\beta_{1}+\beta_{2}{months\_suitable}_{it}+\beta_{3}{REGIC}_{it}+\beta_{4}{urban}_{it}+{\beta_{5}{prior\_outbreak}_{it}+f}_{spat}({lon}_{i},{lat}_{i})+f_{time}(t)+f_{int}({lon}_{i},{lat}_{i},t)$$

Where $Y_{it}$, binary outbreak indicator for municipality *i* (*i = 1, ..., 5,560*) in year *t* (*t = 2001, ..., 2020*), is expected to follow a Bernoulli distribution defined by $p_{it}$, the probability of an outbreak. The Bernoulli distribution is a special case of the binomial distribution where the number of trials is equal to 1. $f_{spat}({lon}_{i},{lat}_{i})$ is the spatial smooth field based on the coordinates (${lon}_{i},{lat}_{i}$) of the centroid of municipality *i*, $f_{time}(t)$ is the temporal smooth function applied to year *t* and $f_{int}({lon}_{i},{lat}_{i},t)$ is the spatio-temporal interaction term. This model is a type of structured additive regression (STAR) model which allows for Bayesian interpretations of additive models by specifying prior beliefs on the smooth functions (8,9). Inference was performed using an empirical Bayesian approach with estimates calculated using restricted maximum likelihood (REML), an approach that has been shown to give more stable estimates than generalised cross validation (10), and more accurate estimates than a full Bayesian approach for binomial models (9). We used the mgcv package in R (8) to fit the spatio-temporal models and to simulate from the posterior distributions of the coefficients to produce mean estimates and 95% credible intervals. Model parameter estimates were compared to an alternative model fitted using an outbreak indicator of over 100 cases per 100,000 residents (considered medium risk by the Brazilian Ministry of Health (4)) to check the results were robust to the outbreak definition.
