## Supplementary table 1 for "The impact of climate suitability, urbanisation, and connectivity on the expansion of dengue in 21st century Brazil"

**Table S1: Distribution of municipalities at each level of influence in the urban network, 2007 (1) and 2018 (2).** The number of municipalities classified as metropoles (largest cities in Brazil, connected throughout the entire country), regional capitals (large cities connected regionally and to metropoles), sub-regional capitals (cities connected locally and to the three largest metropoles), zone centres (smaller cities generally connected only to their neighbours), and local centres (smallest cities typically disconnected from the urban network).

| **Region** | **Metropolis** | | **Regional capital** | | **Sub-regional capital** | | **Zone Centre** | | **Local Centre** | |
| --- | --- | --- | --- | --- | --- | --- | --- | --- | --- | --- |
|  | **2007** | **2018** | **2007** | **2018** | **2007** | **2018** | **2007** | **2018** | **2007** | **2018** |
| **Brazil** | **203** | **214** | **259** | **308** | **270** | **496** | **658** | **437** | **4170** | **4105** |
| North | 5 | 5 | 10 | 13 | 19 | 28 | 38 | 27 | 377 | 376 |
| Northeast | 33 | 33 | 59 | 62 | 57 | 97 | 179 | 140 | 1464 | 1460 |
| Central-West | 25 | 24 | 4 | 7 | 9 | 46 | 83 | 51 | 342 | 335 |
| Southeast | 92 | 95 | 129 | 149 | 104 | 200 | 209 | 120 | 1134 | 1104 |
| South | 48 | 57 | 57 | 77 | 81 | 125 | 149 | 99 | 853 | 830 |
