## Supplementary table 2 for "The impact of climate suitability, urbanisation, and connectivity on the expansion of dengue in 21st century Brazil"

**Table S2: Posterior mean and 95% credible interval (CI) estimates for linear effect parameters, calculated using an outbreak threshold of 100 cases per 100,000 residents, shown on the adjusted odds ratio (aOR) scale.**

| **Coefficient** | **aOR (95% CI)** | | |
| --- | --- | --- | --- |
|  | **Medium risk model^1^** | **Aedes aegypti model^2^** | **Prior outbreak model^3^** |
| Urbanisation | 2.96 (2.66, 3.30) | 3.21 (2.80, 3.65) | 3.83 (3.38, 4.34) |
| REGIC level: metropolis | 1.65 (1.44, 1.88) | 1.38 (1.20, 1.58) | 1.45 (1.26, 1.69) |
| REGIC level: regional capital | 1.77 (1.63 1.92) | 1.51 (1.38, 1.66) | 1.57 (1.44, 1.71) |
| REGIC level: sub-regional centre | 1.42 (1.33, 1.51) | 1.23 (1.14, 1.34) | 1.28 (1.19, 1.38) |
| REGIC level: zone centre | 1.33 (1.26, 1.41) | 1.24 (1.16, 1.31) | 1.26 (1.18, 1.35) |
| Prior outbreak: yes | 2.42 (2.30, 2.55) | 2.01 (1.91, 2.12) | 1.18 (1.13, 1.22) |
| Months with suitable temperature | 1.29 (1.20, 1.37) | 1.34 (1.28, 1.40) | 1.49 (1.38, 1.60) |

^1^ Response variable is dengue outbreak defined as over 100 cases per 100,000 inhabitants

^2^ Temperature suitability set to *Aedes aegypti* limits, between 17.8° and 34.5°C

^3^ Prior outbreak indicator takes account of previous year only
