## Supplementary figures for "The impact of climate suitability, urbanisation, and connectivity on the expansion of dengue in 21st century Brazil"

**a)** **b)**

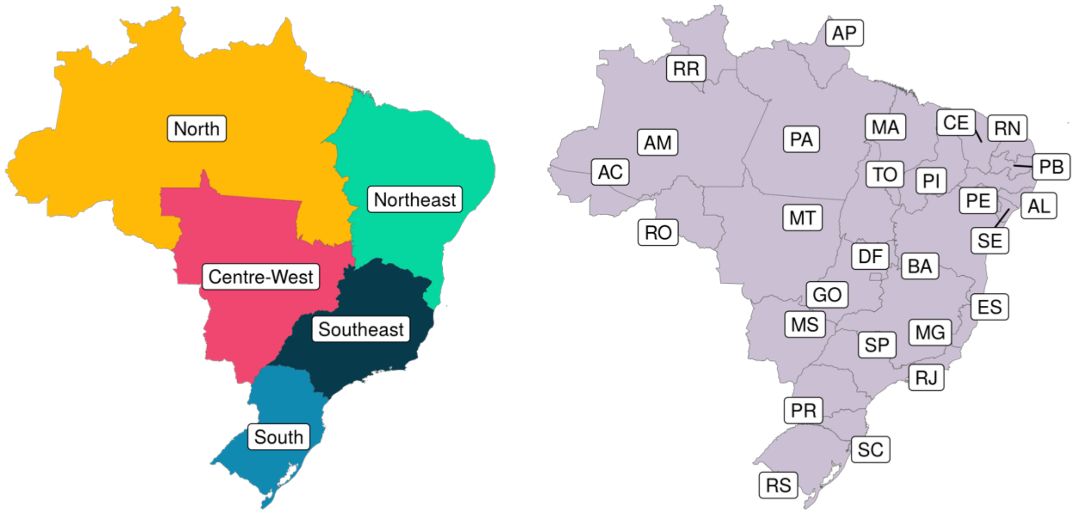


**Fig S1: The organisation of Brazil into a) 5 geo-political regions, and b) 27 federal units**. Abbreviations: AC = Acre, AL = Alagoas, AP = Amapá, AM = Amazonas, BA = Bahia, CE = Ceará, DF = Distrito Federal, ES = Espírito Santo, GO = Goiás, MA = Maranhão, MT = Mato Grosso, MS = Mato Grosso do Sul, MG = Minas Gerais, PA = Pará, PB = Paraíba, PR = Paraná, PR = Pernambuco, PI = Piauí, RJ = Rio de Janeiro, RN = Rio Grande do Norte, RS = Rio Grande do Sul, RO = Rondônia, RR = Roraima, SC = Santa Catarina, SP = São Paulo, SE = Sergipe, TO = Tocantins.

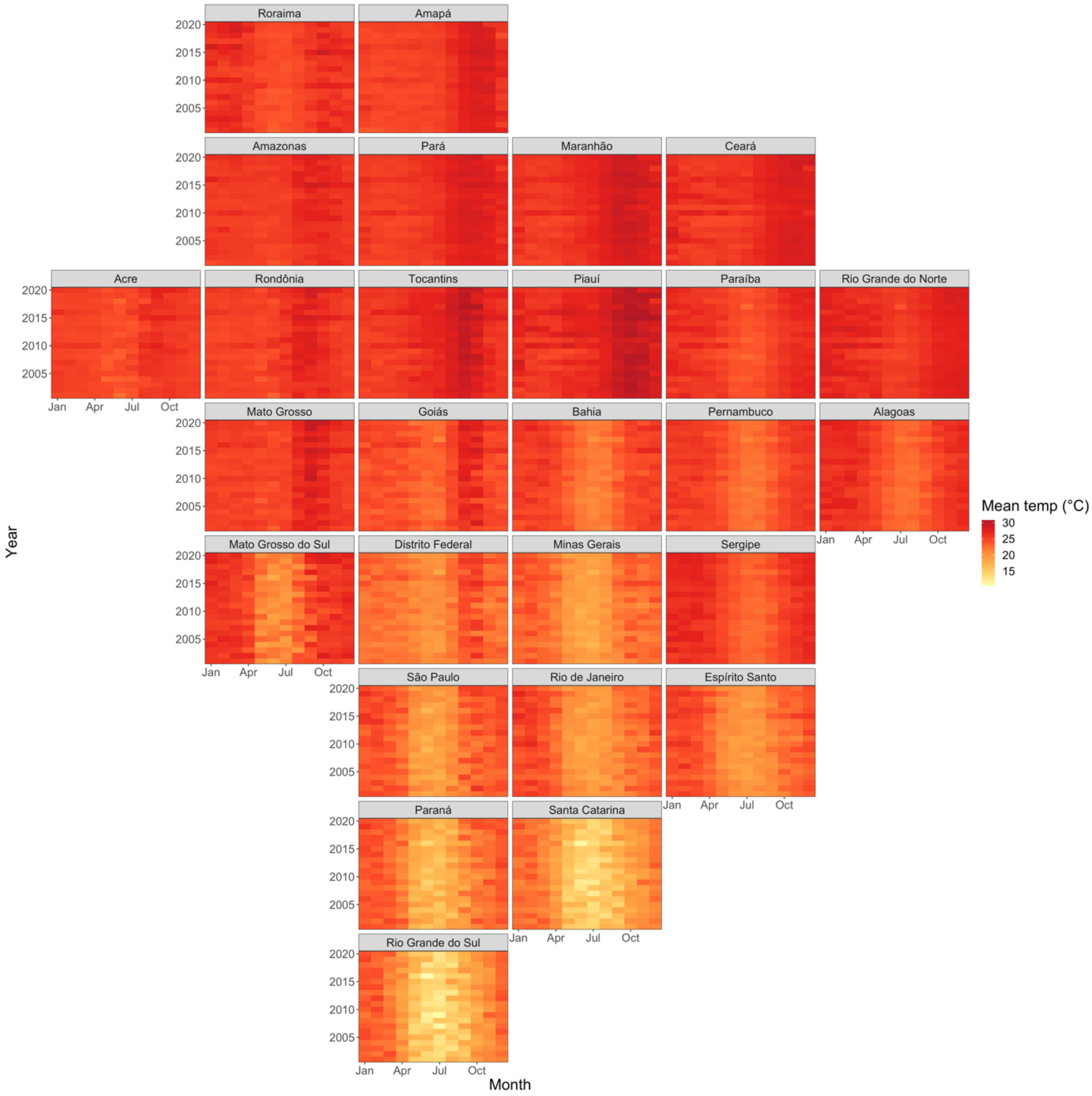


**Fig S2: Average monthly mean temperature (°C) in each Brazilian state January 2001 - December 2020.**

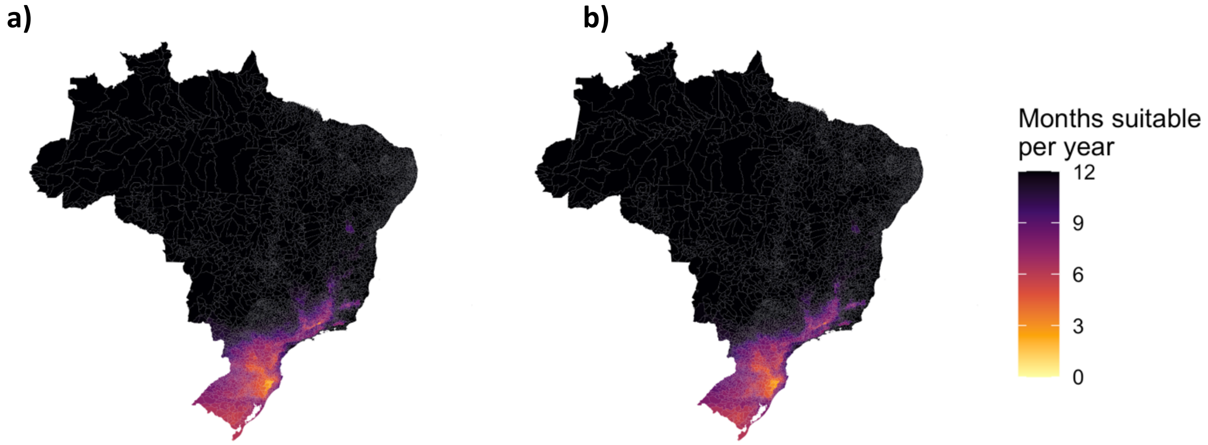


**Fig S3: The average number of months suitable for dengue transmission per year a) 2001 - 2010, and b) 2011 - 2020.** The average number of months with mean temperature between 16.2 and 34.5°C aggregated to the two decades of data. Most of Brazil experiences suitable temperatures year-round apart from areas of South Brazil and areas of high altitude in the Southeast which experience cool winters.

a) b)

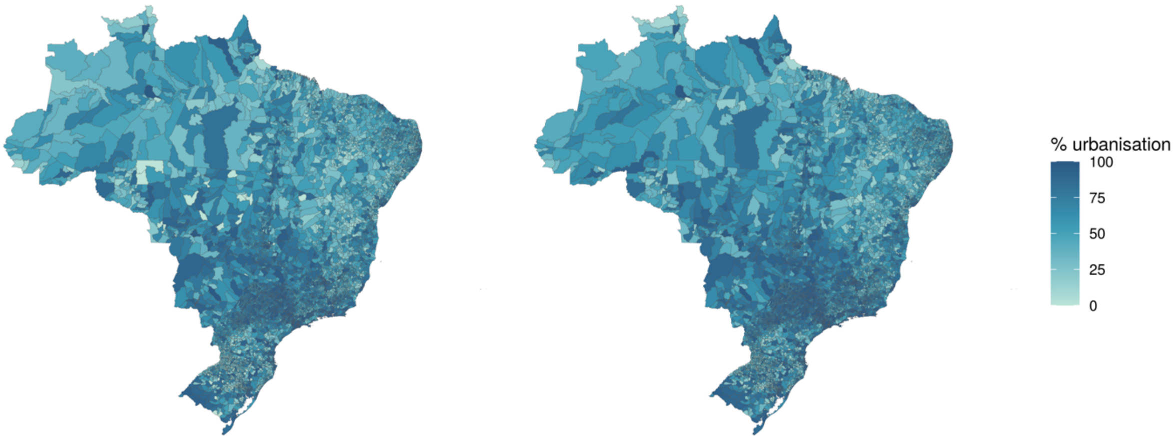


**Fig S4: The percentage of residents living in urban areas of each municipality from the 2000 (a) and 2010 (b) censuses.** Levels of urbanisation differ greatly across Brazil, with the majority of Southeast and South Brazil living in urban areas in comparison to the North and Northeast which has a larger rural population.

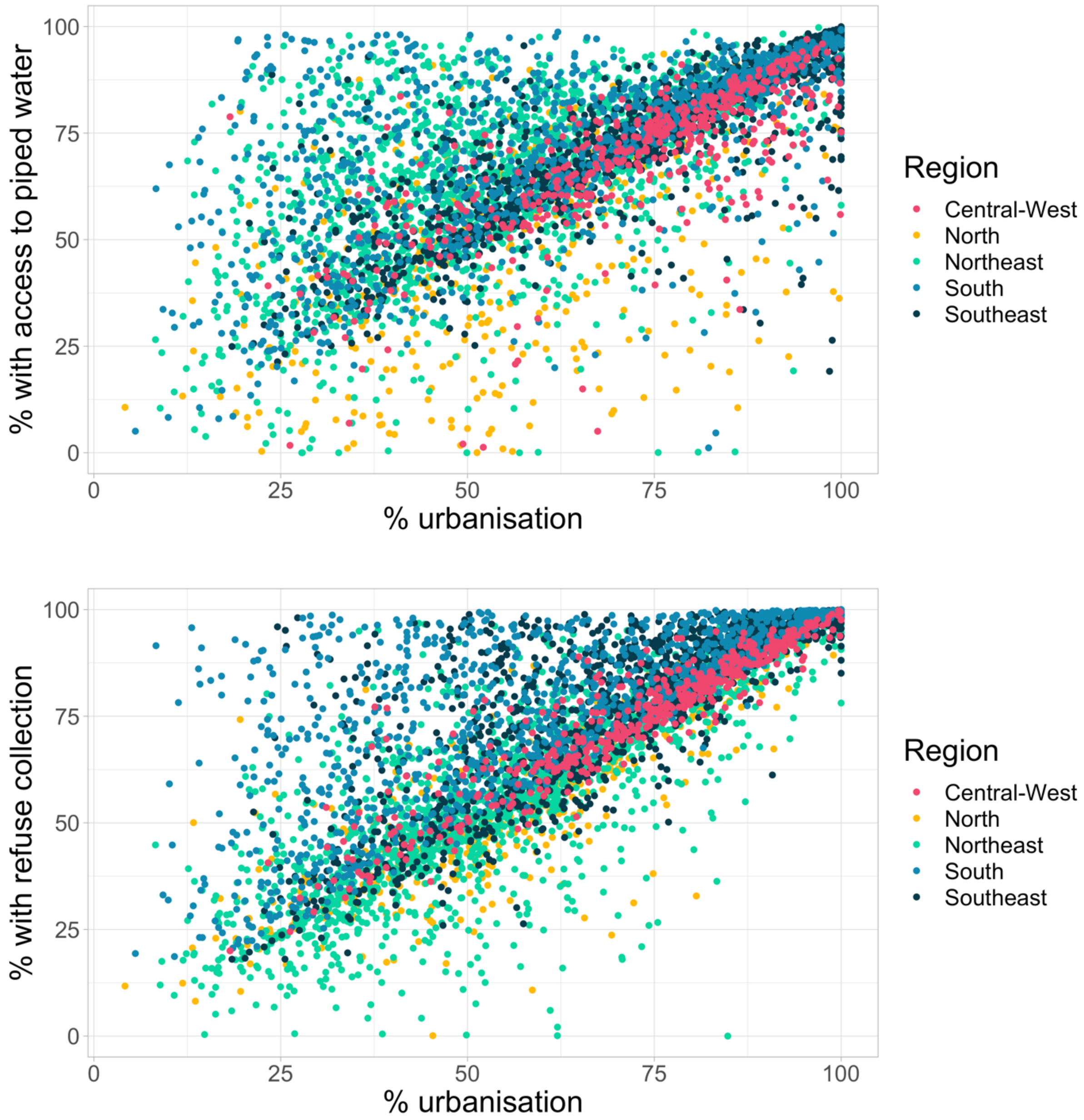


**Fig S5: Scatterplot comparing the percentage of residents with access to piped water (top) and refuse collection (bottom) to the percentage living in urban areas from the 2010 census.** Access to basic services was highly correlated to the level of urbanisation: highly urban areas had highest access to piped water and refuse collection.

a) b)

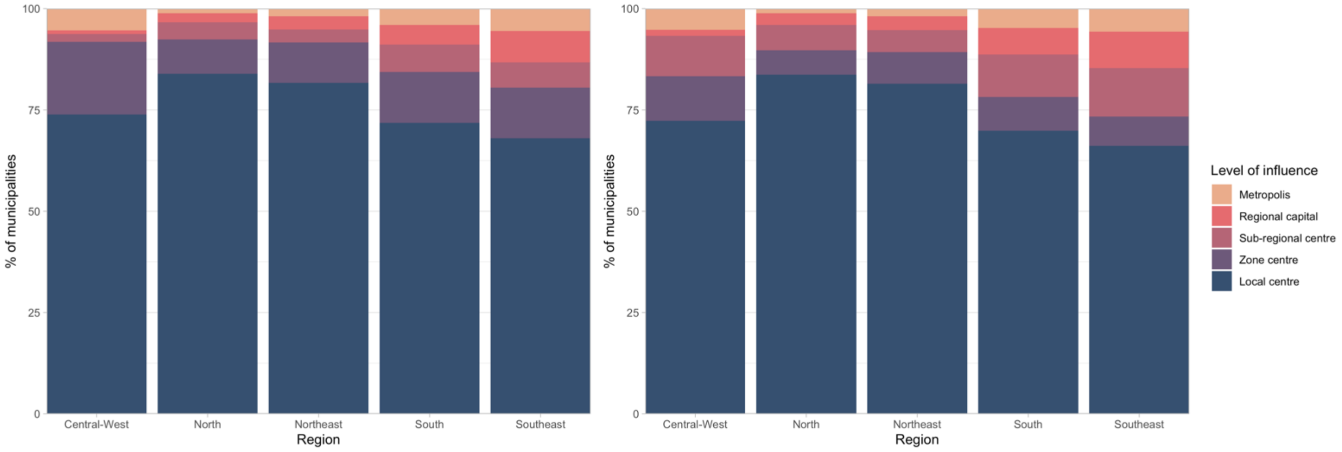


**Fig S6: The proportion of cities in each region at each level of influence in the a) 2007 and b) 2018 REGIC study.** The proportion of high-level cities has increased across the country but the North and Northeast still have noticeably less well-connected cities than other regions. The Southeast and South are by far the most connected regions.

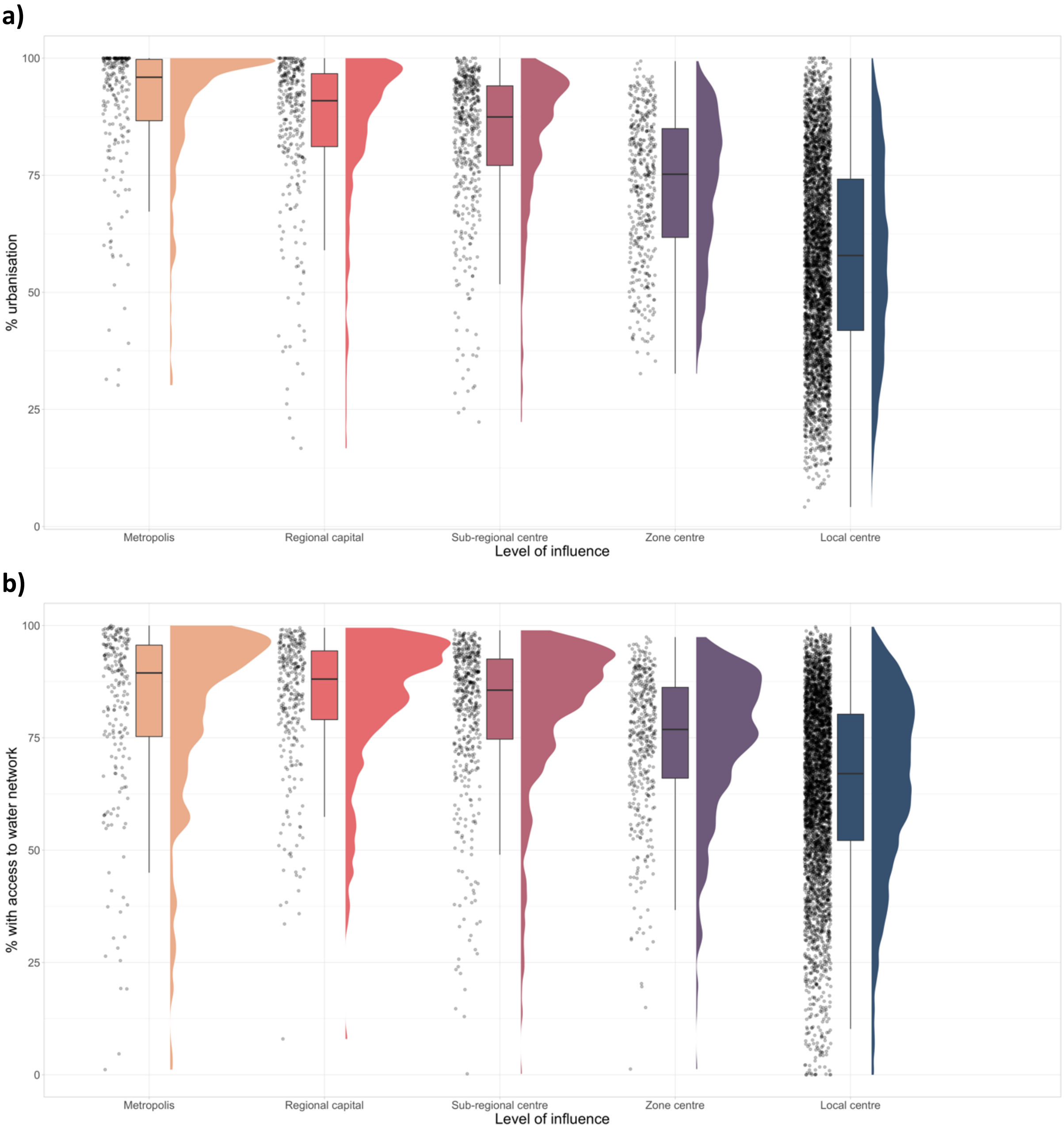
a)
% urbanisation
100
75
50
25
0
Metropolis
Regional capital
Sub-regional centre
Zone centre
Local centre
Level of influence
b)
% with access to water network
100
75
50
25
0
Metropolis
Regional capital
Sub-regional centre
Zone centre
Local centre
Level of influence

**c)**

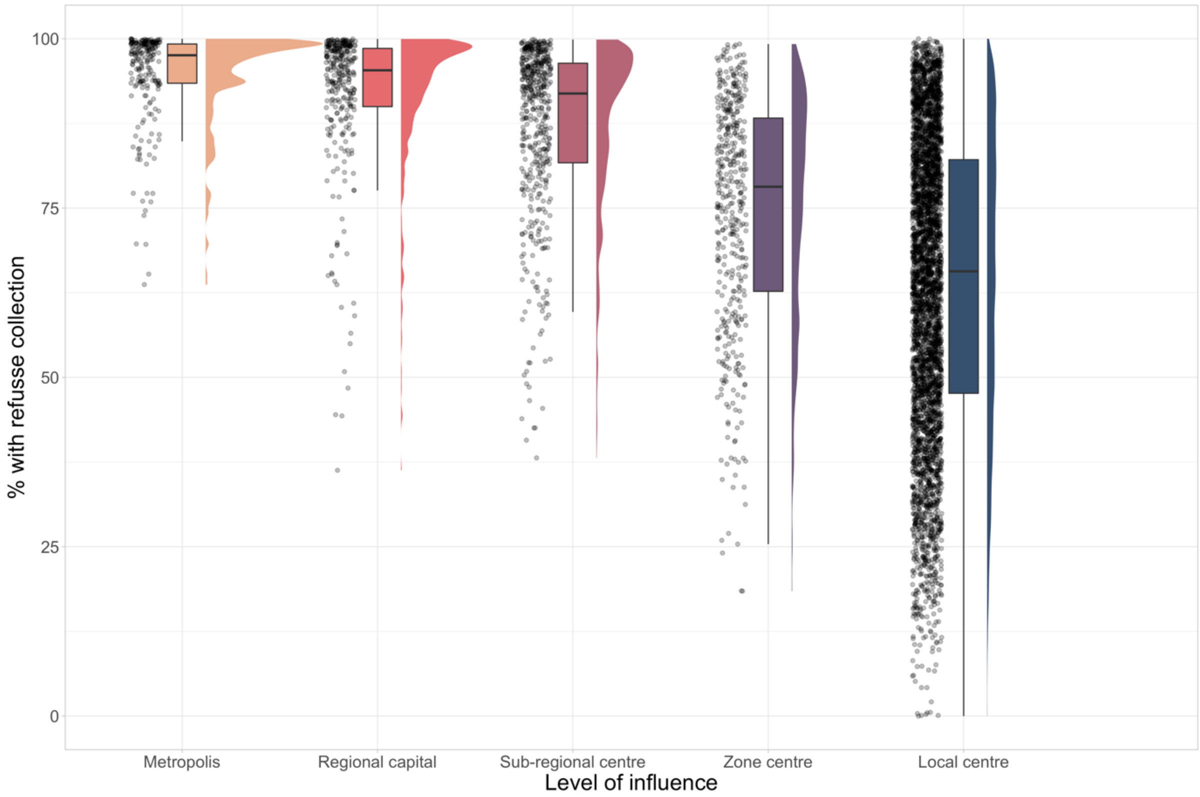


**Fig S7: Raincloud plots exploring the relationship between REGIC level of influence and a) urbanisation, b) access to piped water, and c) refuse collection.** Metropoles and regional capitals have higher levels of urbanisation and access to basic services than municipalities that had lower levels of connectivity within the urban network. Local centres were more varied in terms of basic services and urban levels than the other levels and covered a wide range of city types.

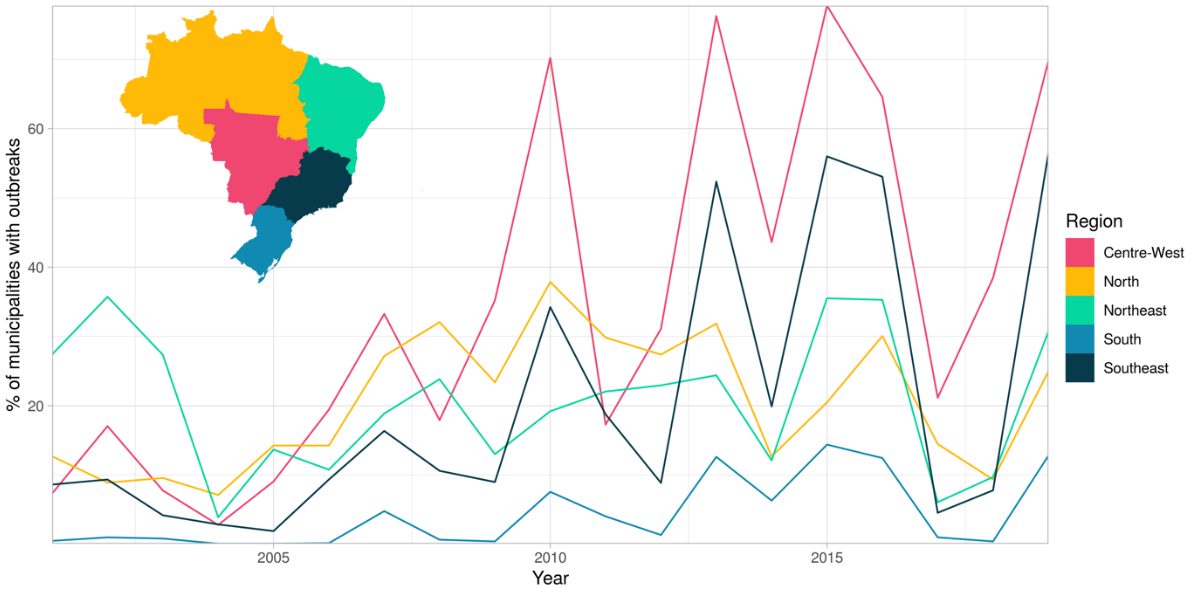


**Fig S8: The proportion of municipalities in each region of Brazil experiencing an outbreak per year 2001 - 2020.** The proportion of municipalities affected by outbreak has increased since 2010 in every region of the country, although outbreaks in South Brazil are still focused on a small part of the region.

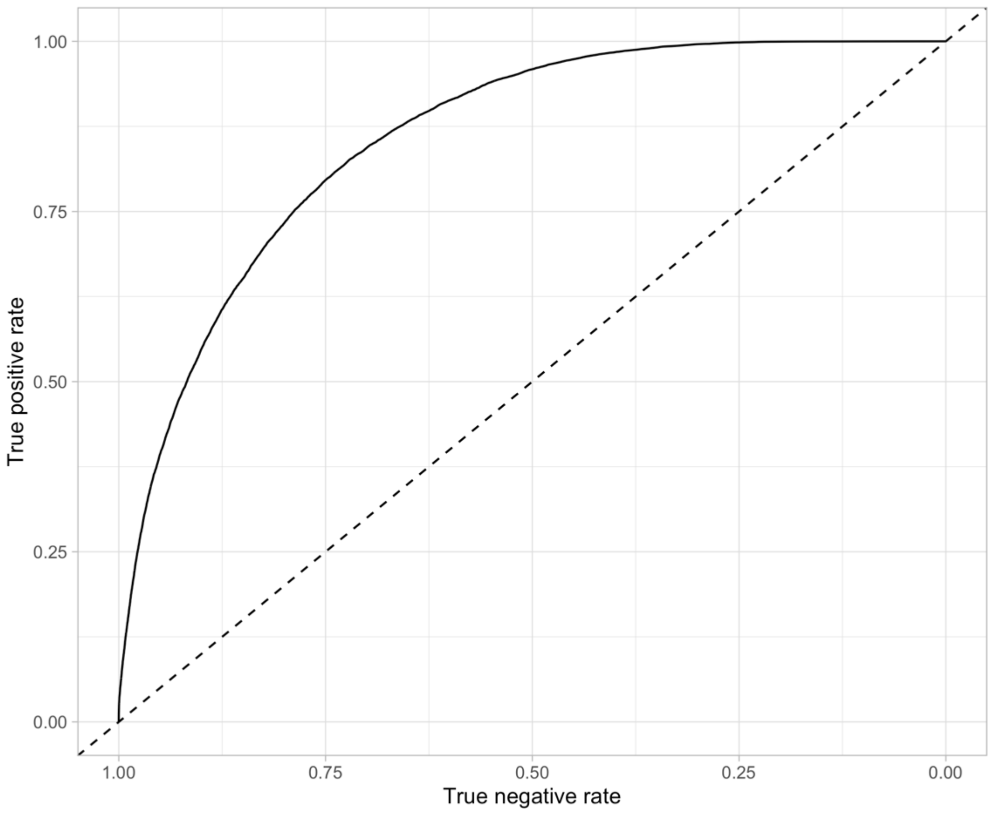


**Fig S9: Receiver operating characteristic (ROC) curve for the final model (solid line) compared to chance (dashed line).** The closer to the top-left corner, the better the predictive ability of a model. As the ROC curve lies above the dashed reference line, this model performs better than chance.

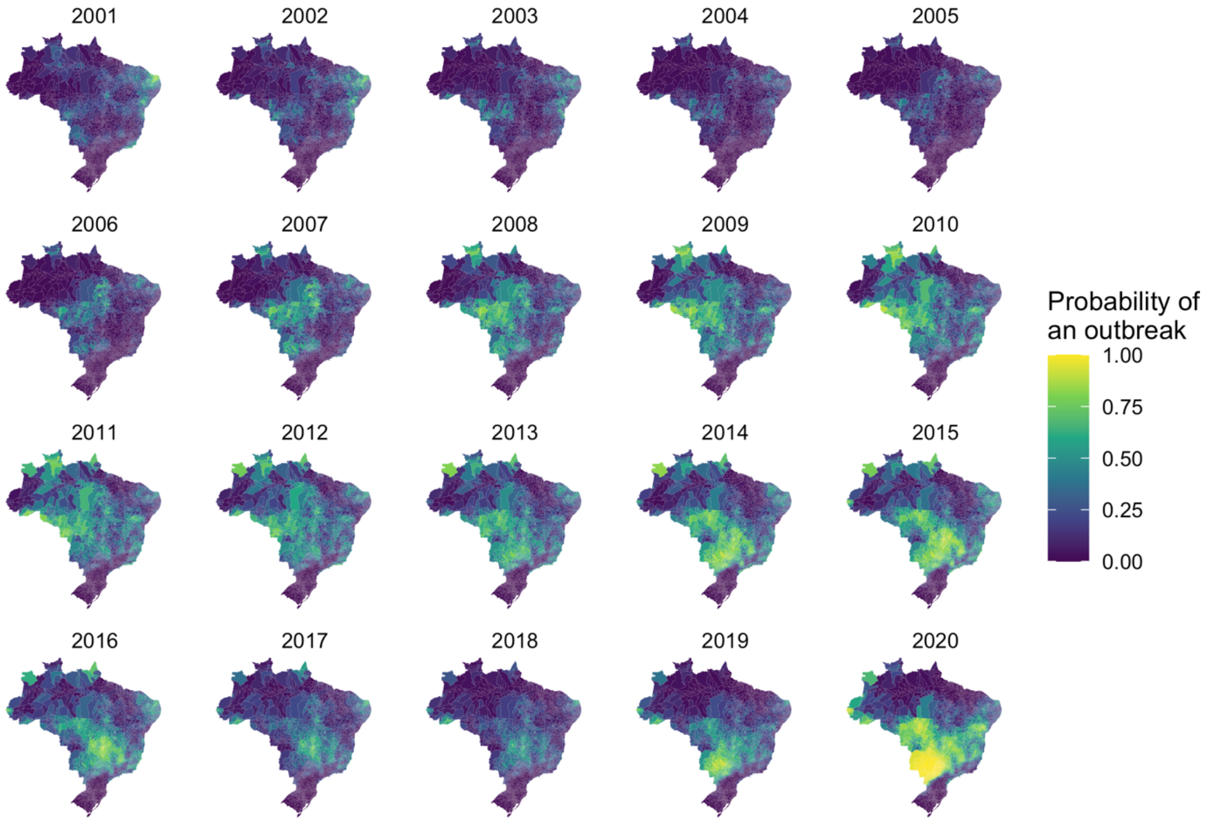


**Fig S10: The probability of an outbreak estimated from the model for each year 2001 - 2020.** The mean probability of an outbreak estimated by taking 1000 simulations from the posterior distribution of the response and transforming the outcome using a probit function

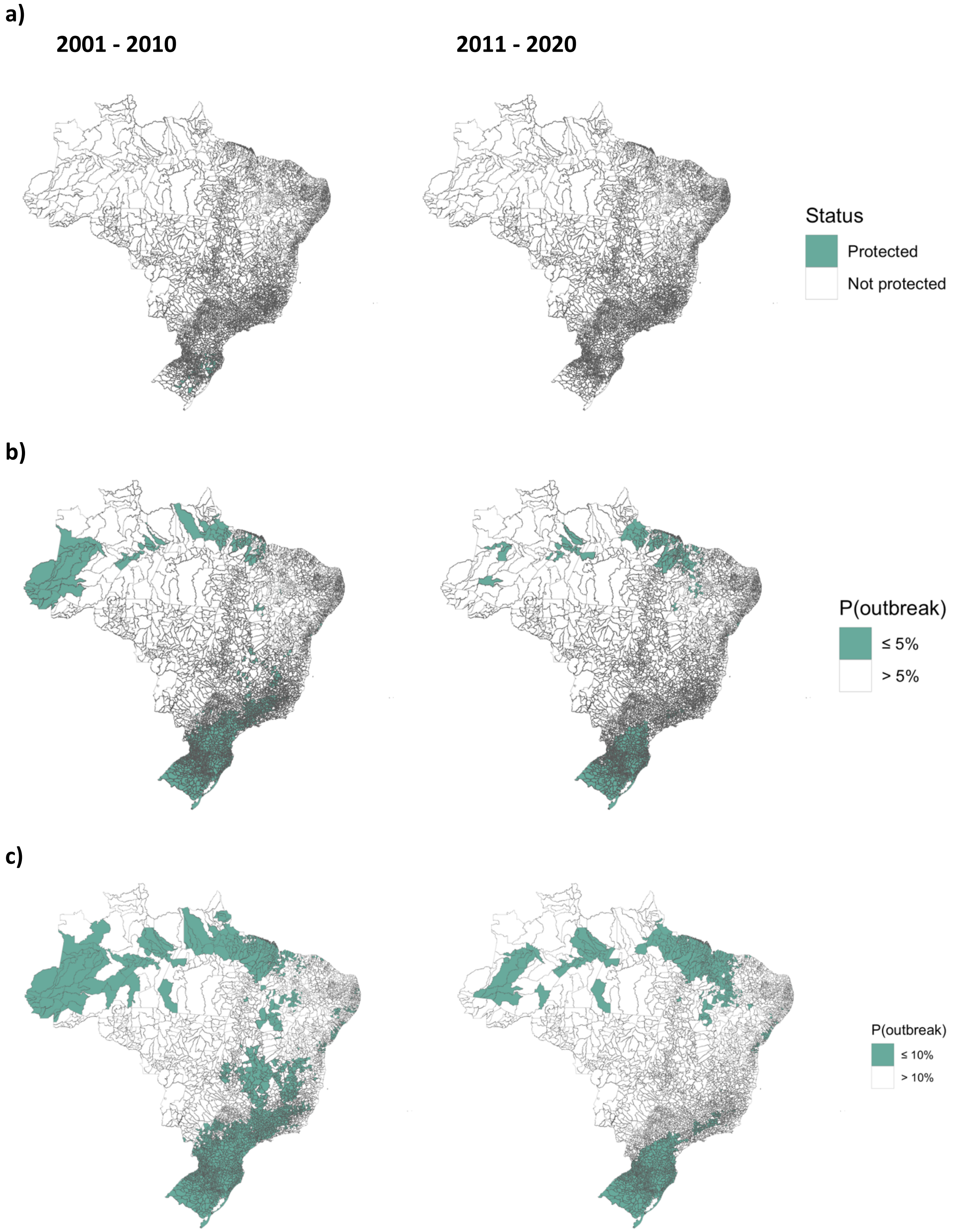
a)
2001 - 2010
2011 - 2020
Status
Protected
Not protected
b)
P(outbreak)
≤ 5%
> 5%
c)
P(outbreak)
≤ 10%
> 10%

**d)**

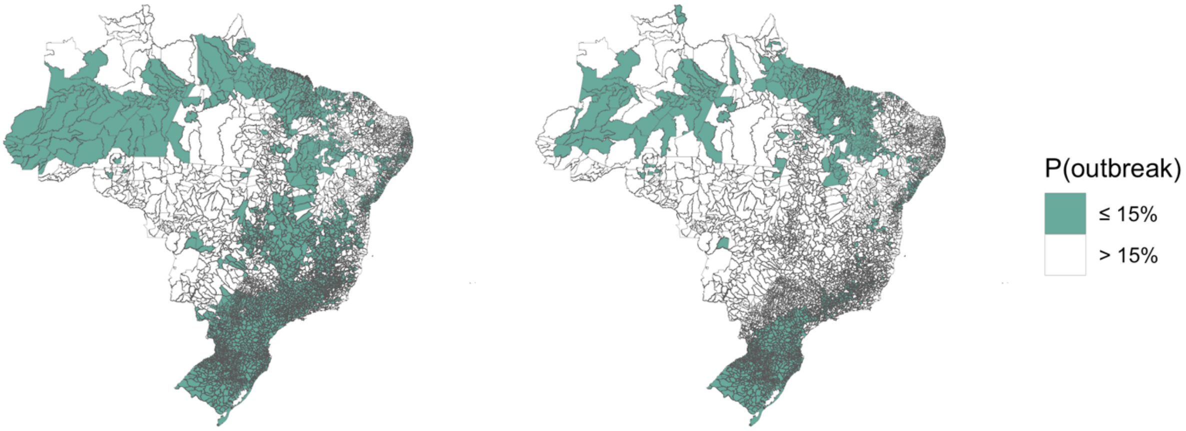


**Fig S11: Comparison of different risk thresholds to define current geographical barriers to dengue outbreaks.** Municipalities were considered 'protected' if the probability of an outbreak was less than or equal to the threshold a) 0%, b) 5%, c) 10% or d) 15%. The threshold of 10% was chosen as it was the most comparable with previous studies.
